# Do Faster-Trained Physicians Fill the Gaps? Geographic Concentration of Emergency Medicine physicians with different postgraduate training in Ontario Canada

**DOI:** 10.1101/2025.04.29.25326528

**Authors:** David Kanter-Eivin, Calvin Armstrong, Anil Esleben, Grant Sweeny, Michaela Dowling, Asil El Galad, Stephenson Strobel

## Abstract

**Background:** Emergency departments in underserved areas face chronic staffing challenges. One possible solution is to use physicians who are quicker to train and more pervasive in lieu of more extensively trained physicians. Canada allows for emergency medicine specialization via a 3-year pathway (CCFP(EM)) and a 5-year pathway (FRCPC) which means different geographic areas are exposed to EM physicians with different training lengths.

**Methods:** We examine Ontario, Canada which has both widespread geographic diversity and emergency providers with these two lengths of postgraduate training. We scrape the College of Physicians and Surgeons of Ontario public registry in 2015 and 2024. We map the geographic distribution of physician types and estimate spatial autocorrelation measures using global and local Morans I to determine whether these physicians became more geographically concentrated.

**Results:** Between 2015 and 2024, the number of CCFP(EM) and FRCPC physicians increased in overall numbers but their unique locations remained stable. Mapping of these locations suggests clustering into urban or suburban areas in the province. CCFP(EM) physicians have become more concentrated over time (Morans I of 0.234 and 0.308 in 2015 and 2024) relative to FRCPC physicians (Morans I of 0.096 and 0.103).

**Conclusion:** We find that, from 2015 to 2024, emergency physicians have become more concentrated in the province of Ontario due to CCFP(EM) physicians concentrating around urban areas with academic centres. Policies relying on less extensively trained providers to plug staffing gaps may not necessarily be effective in improving equitable access to physicians.

**Note: Preprint:** This preprint reports new research that has not been certified by peer review and should not be used to guide clinical practice.

## 1. Background

Rural areas in North America face a physician workforce shortage problem that undermines equitable access to emergency medical care. Staffing shortages are one of the primary contributing factors to emergency department (ED) overcrowding^1^ and these disproportionately affect rural EDs. In Canada in 2016, nearly 40% of EDs in urban settings and 62.5% in rural settings were insufficiently staffed^2^, and in 2023, there were 1,184 ED or urgent care closures in Ontario with most occurring in smaller, rural centers^3^. Staffing is expected to worsen in Canada, with a projected shortage of 1,500 certified emergency physicians by 2027^2^. In the United States, the workforce in rural ED settings has also declined relative to population^4^. The consequent ED closures increase strain on surrounding health-care facilities^5^, cause longer transportation times^6^, and increase burden on medical transportation services^7^. Such closures also pose an issue for non-emergent patients in rural areas who rely on local EDs for their primary care needs^8^.

To provide more equitable access to emergency care, one solution that has evolved has been substituting delivery by differently-trained healthcare providers. This occurs in many rural areas in the United States where family physicians and physician extenders compose a significant percentage of care providers^9^. In Canada, the presence of an alternate shorter training program for emergency physicians, the three year CCFP(EM) training path, effectively reduces the length of training that emergency physicians undergo, and substitutes these more common providers in lieu of more extensively trained physicians who train for five years through the FRCPC training program. While this may increase the number of physicians able to provide emergency medicine services, this strategy will only improve access to care if physicians with less extensive training remain in underserved areas. If these physicians are drawn to already well-served areas, this strategy may not produce more equitable care.

We examine the geographic concentration of emergency physicians in Ontario, Canada. Ontario has two features that are useful for our research. First, it is geographically diverse: it includes urbanized environments with tertiary care centers, rural areas where patients must drive to obtain community ED care, and remote areas in the Northwest of the province that are often only accessible by air travel and deliver care through nursing stations. To emphasize how large an area it is, the furthest points of the province are approximately 1,800 kilometers apart.

Second, emergency medicine residency training in Canada proceeds along two distinct streams: the five-year FRCPC residency program in which residents and eventual graduates are usually based out of large tertiary care centers, and a two-year family medicine residency with an additional specific year of emergency medicine training which is the CCFP(EM) training path. This latter program produces emergency physicians who more commonly work in community settings, including rural and remote areas^10^. These two aspects allow us to evaluate whether the concentration of more or less extensively trained emergency physicians has changed over time.

We leverage a scraped data set of all physicians in Ontario that have ever been registered with the College of Physicians and Surgeons of Ontario (CPSO), including those who completed these two types of emergency medicine certifications. These data were scraped in 2015 and in 2024, allowing us to examine the evolution of the geographic concentration of these more and less extensively trained specialty physicians. We estimate geospatial concentration of these two specialties by measuring spatial autocorrelation for both types of emergency medicine providers, as well as other medical specialties for comparison, in each year we have data. We provide two novel contributions. The first is technical: we apply measures of spatial concentration to evaluate distributions of emergency physi-cians. The second is relevant to policy: we estimate whether the concentration of these two differently trained emergency physicians have changed over time. Empirical understanding of how differently trained emergency medicine specialists might concentrate is important where policymakers rely on less extensively trained providers to provide care in underserved areas.

## 2 . Methods

### 2.1. Institutional Background: Emergency Medicine Training in Canada

Before 1980, almost all emergency medical care in Canada was delivered by general practitioners who completed one-year internships. In many cases, overnight shifts were covered by residents without supervision by an attending physician^10^. During this time, policymakers became concerned about the quality of emergency care provided from physicians not specifically trained in emergency medicine. A general policy framework was agreed that physicians should undergo additional specialized training to practice emergency care; however, the specifics of the policy resulted in a turf war over training emergency medicine specialists^11^,^12^. Canadian postgraduate medical education was, and continues to be, split between two training bodies: the Royal College of Physicians and Surgeons of Canada (RCPSC) and the College of Family Physicians of Canada (CFPC)^10^,^12^,^13^. During the 1970s, there was a contentious debate over which certifying body should be responsible for training emergency physi-cians: both the RCPSC and CFPC sought this responsibility. The former trains specialist physicians whereas the latter trains generalist, family physicians. The CFPC also certifies additional training for family medicine specialists in knowledge areas particularly relevant to generalist care such as palliative care, obstetrics, and emergency medicine.

As a compromise, both colleges created training programs. The RCPSC developed a four-year program which then became a five-year program. The CFPC developed a program that was an additional year of training that commenced after a two-year family medicine residency. The original implementation required that both colleges jointly certify physicians who completed either stream. However, in 1982 the colleges could not agree with respect to certification and “agreed to disagree”, resulting in two separately-certified streams with differing training lengths^12^.

The importance of this policy context is that these parallel training streams led to two differently trained emergency physician professionals. RCPSC emergency medicine specialists, designated as FRCPC, have 5-years of postgraduate training in emergency medicine, usually work in academic centers and treat more acute patients. CFPC emergency medicine specialists, designated as CCFP(EM), have a shortened training program with more experience in family medicine. There is a higher annual volume of graduates from these CCFP(EM) programs and they tend to work in community settings.

### 2.2. Data

We use a dataset of all physicians in Ontario that have ever been registered with the CPSO. This data is directly webscraped from the CPSO public registry and contains information on specialty training and six digit postal codes that correspond to a practice location. These data are complete for all physicians who are actively practicing in the province as of the date of scraping. We restrict our sample of physicians to those who practice independently. These data were scraped twice: once in 2015 and again in 2024 which allows us to estimate changes in our measures of interest between these years.

We aggregate these individual, physician level data to two geographies which are our unit of analysis. In our main analyses, we aggregate the numbers of each physician type within an area to a 10 by 10 km tessellated square area within Ontario. In our descriptive statistics, we aggregate to a three digit postal code or forward sortation area (FSA) level because detailed data on population levels are only available at this level^14^. This aggregation gives counts of the CFPC and RCPSC emergency medicine specialists who primarily work in each 10 by 10 km area or three digit postal code. In order to identify whether our results are driven by secular trends, we compare Moran’s I estimates for EM specialties to other groups of physicians, including internal medicine physicians, general surgeons, and family physicians. This is in part to see whether underlying trends that affect the whole province (and so should affect the concentration of other physicians) are occurring. An example of such secular trends would be population growth in cities or new hospitals opening that draw in all types of physi-cians^15^. For the cases of internal medicine physicians and general surgeons, individuals would have completed a five year residency through the Royal College of Physicians and Surgeons of Canada. In the case of family physicians, individuals would have completed a two year residency through the College of Physicians and Surgeons of Canada.

We use two additional sets of data for summary statistics. These are census profiles at the FSA level for the 2016 and 2021 census of Canada^14^. We map our physician counts in each FSA in 2015 and 2024 to data on FSAs in the closest available years. We demonstrate summary statistics for these FSAs to demonstrate the types of geographic areas EM physicians are located.

We calculate spatial autocorrelation using Moran’s I to evaluate geographic concentration patterns^16^.

### 2.3 Estimates of spatial and geographic concentration of physicians

Our interest is in estimating a measure of distribution and concentration of the two emergency medicine specialties in each year that we observe them. To do so, we take the centerpoint of the six-digit postal code in which the physician locates their primary practice. We rasterize this as a set of points throughout the province. We then plot these across a provincial map. For our main analysis, we do not attach these physician practice locations to their specific geographic postal code, but rather a set of geographic units of 10 square kilometers. The rationale for this lies within the large variation in the size of the postal codes in Ontario: in places like Toronto, these can be as small as a city block, whereas in northwestern Ontario they become as large in area as some US states. This inconsistency creates a variant of the modifiable areal unit problem where, within units, we are aggregating to vastly different geographic areas. To avoid this issue, we use a tessellated map which splits Ontario into 8,964 units that are approximately 10 square kilometers^17^. We aggregate physician counts to this 10 by 10 km unit level.

As a first descriptive exercise, we examine the raw counts of physicians within the province and plot their locations throughout the province. We create counts for each physician type within each of our geographic units for each year of observation. There are physicians who have multiple specialties (for example, physicians completing a family medicine residency and then an internal medicine residency), so these individuals are counted in each physician group. We map the locations of these physicians.

We then estimate geospatial concentration of physician types within the province of Ontario over time. We estimate a Moran’s I which quantifies the degree of spatial clustering, and is often used in hot-spot analysis where researchers are interested in identifying areas of high prevalence of disease like cancer^16^. For our main results we assume a distance weighting matrix such that only other geographical units within 19 kilometers are included. We also provide additional estimates using 38 and 96 kilometer distance weighting matrices to sensitivity check our results (Supplementary Table e2). We produce separate estimates for the years of 2015 and 2024 and for each physician type in our data. We also estimate Local Moran’s I to provide some evidence on where these areas of concentration are within the province. We generate 999 permutations to calculate a pseudo *p*-value. We use a cutoff of 0.01 for this to mitigate issues with multiple hypothesis testing. The Moran’s I typically ranges from approximately −1 to +1: positive Moran’s I values indicate positive spatial autocorrelation, suggesting greater geospatial clustering of physicians; conversely, negative values suggest a geographic dispersion pattern, i.e. tessellated blocks with high numbers of physicians are located near other tessellated blocks with high numbers of physicians.

Finally, as an adjunct measure to our Moran’s I we estimate a GINI coefficient and plot a Lorenz curve of geographic inequality of access to CCFP(EM) and FRCPC physicians in 2015 and 2024. These are conducted at the tessellated block level. The Lorenz curve plots the cumulative share of physicians against the cumulative share of geographic units, ordered from lowest to highest physician count. We restricted the analysis to areas with at least one physician to focus on within-system disparities. The Gini coefficient, a summary measure of inequality ranging from 0 (perfect equality) to 1 (perfect inequality), was calculated using the rank-share method. This approach computes the Gini as one minus twice the area under the Lorenz curve, estimated by summing the product of each area’s share of physicians and its relative rank (as a share of total areas).

## 3. Results

Over the period from 2015 to 2024, the number of CCFP(EM) physicians licensed in Ontario increased from 1076 to 1399. The number of FRCPC physicians increased from 424 to 599. In Table e1, we show several select summary statistics that are based on values at the FSA level for each year. Similar numbers of FSAs have access to CCFP(EM) physicians whereas slightly more FSAs have access to FRCPC physicians in 2024 than in 2015. FSAs with an emergency medicine specialist tend to have larger populations, and have higher levels of employment. The average age and average income of persons within these FSAs tends to be similar.

Figure 1 and e1 map the spatial distribution of emergency physicians in Ontario. There is a high concentration of all types of these physicians within southern Ontario, with fewer emergency specialists located in the sparsely populated areas of Northwestern Ontario. FRCPC physicians are largely located in larger cities such as Hamilton, Ottawa, and Toronto, and in towns with academic universities such as Kingston, London, and Thunder Bay. There is a larger proportion of CFPC(EM) physicians who work in areas outside of these settings, but these physicians remain clustered in southern Ontario around larger centers.

**Figure 1.**
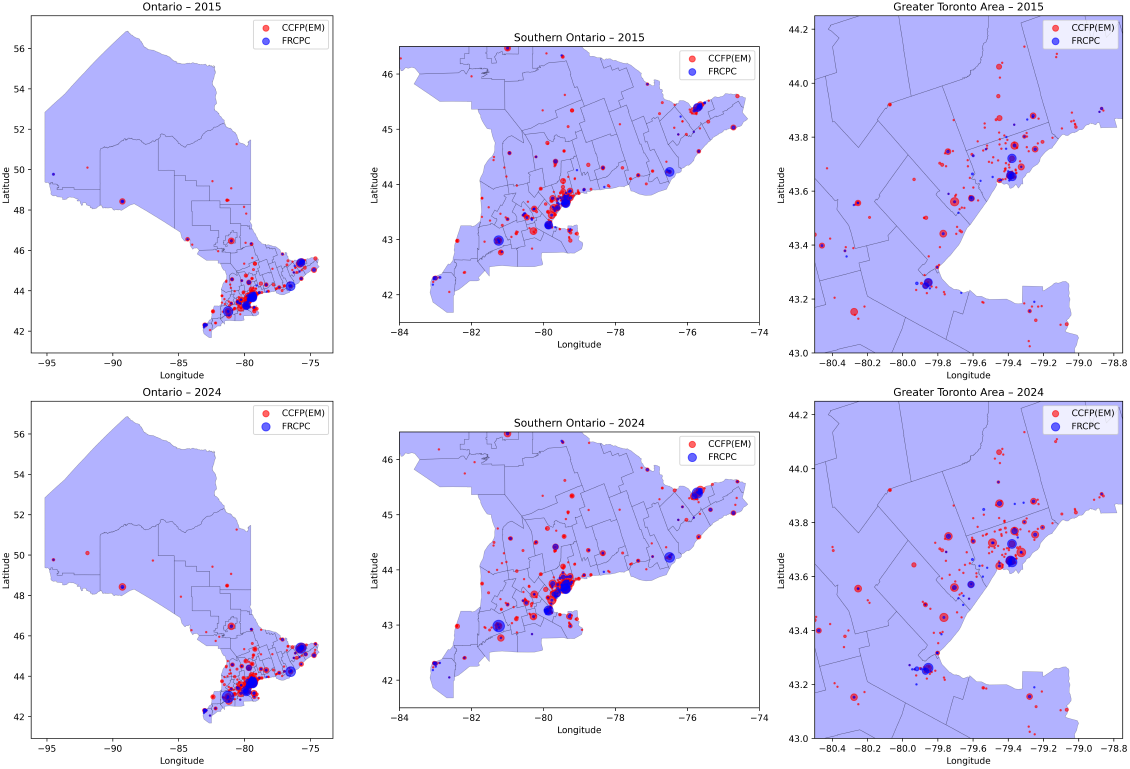
Map of the locations of emergency medicine specialists in Ontario in 2015 and 2024. Bubble size corresponds to the number of physicians in one location.

Figure 2 demonstrates local Moran’s I estimates of spatial clustering. Areas with high clustering (high-high clusters) are indicated with red and areas with low amounts of spatial clustering (low-low clusters) are indicated with blue. Light red is used for areas of high concentration surrounded by areas of low concentration (high-low clusters). High concentration occurs in areas around Toronto in southern Ontario and some other areas like Ottawa in eastern Ontario. There are more hot spots for CCFP(EM) specialists in both 2015 and 2024 relative to FRCPC physicians. This occurs both in a dispersed area around Toronto where the majority of these physicians are located, but also in the more sparsely serviced areas in Northwestern Ontario. Supplementary Figure e2 shows local Morans I estimates continuously without consideration of cluster type (quadrant).

**Figure 2.**
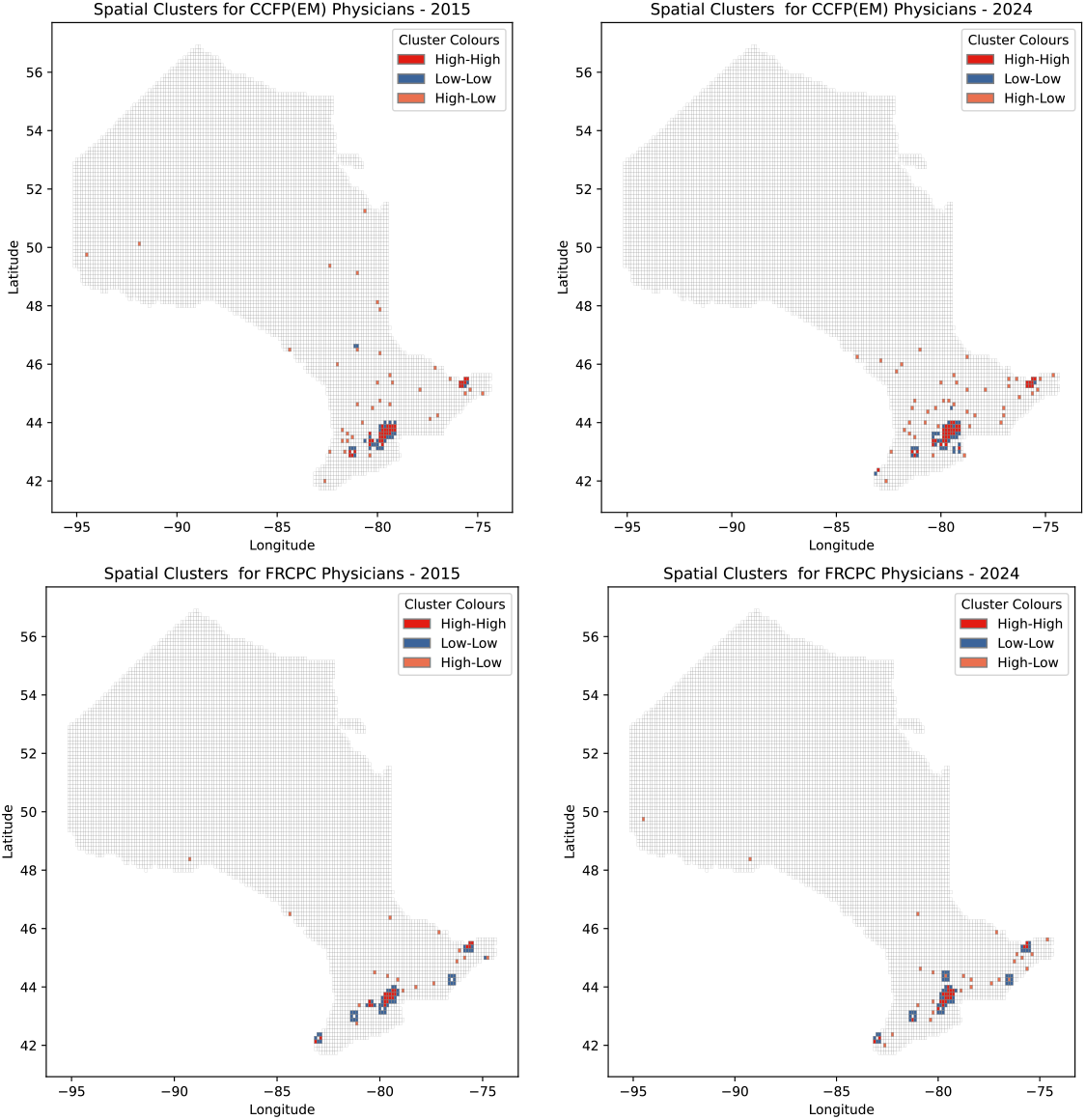
Spatial Autocorrelation Clusters for RCPSC and CFPC specialists using Local Moran’s I. Clustering was considered significant with *p* ≤ 0.001.

Figure 3 demonstrates bar graphs of the overall Moran’s I for the two emergency medicine specialist types in each year. Using a 19×19km distance band, we find an overall Moran’s I for CCFP(EM) physi-cians of 0.234 (*p* ≤ 0.001) in 2015 which increases to 0.308 (*p* ≤ 0.001) in 2024. We find that an overall Moran’s I for RSPSC physicians in 2015 is 0.095 (*p* ≤ 0.001) which remains relatively stable at 0.103 (*p* ≤ 0.001) in 2024. We find that there are similarly minor changes in geographic concentration of general surgeons and internal medicine physicians. We find a somewhat larger increase in the concentration of family medicine specialists. *p*-values and Moran’s I for all specialties, as well as results for 36km, and 96km distance weights, can be found in Supplementary Table e2.

**Figure 3.**
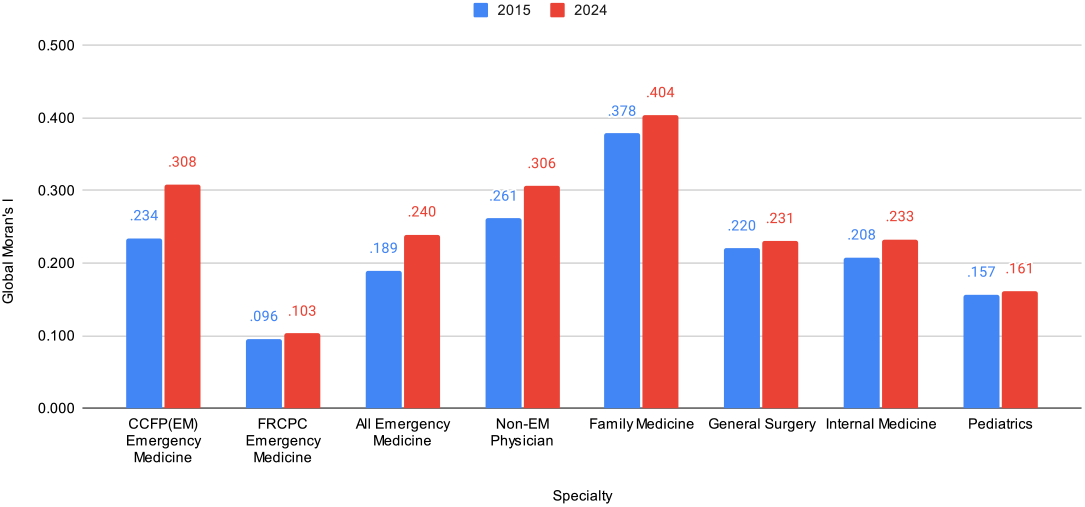
Global Moran’s I estimates for each year and for select specialists. Higher Moran’s I values indicate tighter geographic clustering of physicians.

Figure 4 demonstrates the Lorenz curves for geographic inequality of access to emergency medicine physicians along with estimates of the Gini coefficient in the note of the figure. We find increases in inequality across both types of emergency physician from 2015 to 2024. For CCFP(EM) physicians, this inequality increases from 0.650 to 0.676. For FRCPC physicians, this inequality increases from 0.717 to 0.748. Both Gini estimates suggest very unequal geographic access to emergency physicians at baseline.

**Figure 4.**
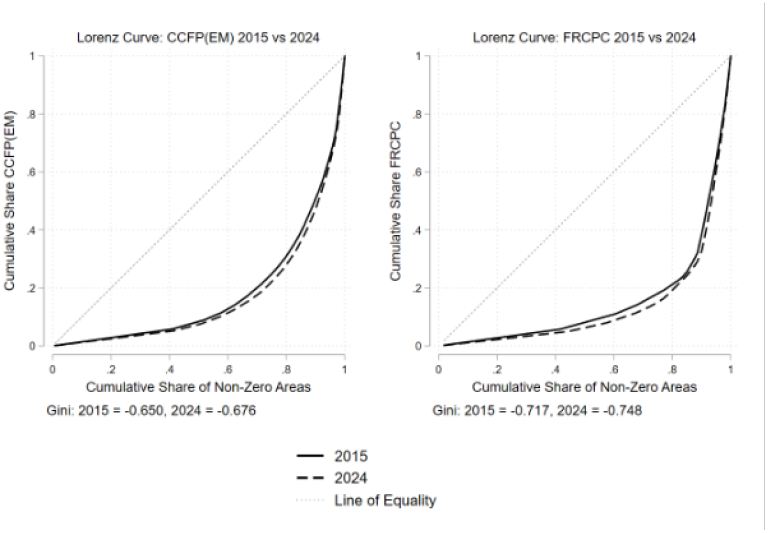
Plots of Lorenz curves for CCPF(EM) and FRCPC physicians for 2015 and 2024. These demonstrate inequality in the numbers of physicians by tessellated block. Perfect geographic equality is denoted by the line of equality. Curves that deviate away from this line indicate increasing inequality in emergency physician access across blocks.

## 4. Discussion

We provide three major insights into the geographic concentration of emergency medicine specialists in Ontario over the period of 2015 to 2024. First, we provide insight into the levels of emergency specialists and their distribution throughout the province. While the number of overall specialists has increased, the number of locations serviced by these physicians has remained relatively stable. Second, we provide evidence that what has occurred over nine years is not increasing overall dispersion through the province. Instead there is geographic concentration around urban areas in Ontario and especially in the Greater Toronto area. Our estimates of Moran’s I, together with the stable number of serviced areas, suggest that the distribution of physicians has become more concentrated in value (i.e., more physicians per location), and also more spatially clustered, particularly in urban cores and suburban areas. Our Lorenz curves and GINI coefficients support this finding in that their measures of inequality have increased during this time for both types of emergency medicine physicians. Finally, we find that our estimates of geographic concentration are not due to more extensively trained, FRCPC physicians geographically concentrating. Instead, we find that less extensively trained, CFPC(EM) emergency physicians are concentrating towards urban areas in the province as evidenced by our local Moran’s I mapping.

Our results support anecdotes that access to an emergency specialist is highly dependent on location within the province^18^,^19^. Our mapping of this data onto 10×10km blocks suggests that both types are concentrated in southern Ontario around the Greater Toronto Area, but there are more CCFP(EM) specialists in remote and rural northwestern Ontario relative to FRCPC specialists. However, we find that over the last nine years, despite an increase in the overall number of both types of specialists, similar areas in Ontario have access to CCFP(EM) trained physicians. We find only a modest increase in access to more extensively trained FRCPC emergency specialists by the number of FSAs in which these physicians are located.

However, what is more concerning is the change in this concentration of emergency physicians over time. Reductions in the geographic prevalence of emergency physicians throughout the province might be tolerable if this came with more dispersion throughout the province. Such a policy of concentrating these specialists in fewer but more geographically widespread hubs might actually improve equity in access to these physicians. However, our estimates of global and local Moran’s I indicate that emergency physicians—especially those with CCFP(EM) training—have become increasingly concentrated in fewer, spatially clustered urban areas. This concentration, combined with rising inequality as measured by the Gini coefficient, suggests a reduction in redundant access: while many areas still have at least one emergency-trained physician, fewer areas have a robust supply. Fewer areas have access to multiple CCFP(EM) trained physicians. This coupled with our previous result on the number of areas where these physicians are located suggests that equity of access to emergency physicians has deteriorated in Ontario over our period of observation. This evidence that emergency medicine trained physicians are increasingly concentrating in highly urbanized environments is consistent with other evidence from the US that suggests similar agglomeration over time^20^.

Importantly, the increase in spatial concentration is driven almost entirely by CCFP(EM) physicians. FRCPC-trained emergency specialists show a relatively stable geographic footprint over time, with little change in Moran’s I. This divergence suggests that the centralization trend is not generalizable across emergency medicine, but is specific to the subset of less extensively trained physicians. We observe similarly modest changes in the concentration of other specialists over time such as pediatricians. This increase in concentration among CCFP(EM) specialists is not, then, the result of a secular trend that affects all specialties. The only similar increase in concentration that we observe occurs among family physicians, but this is smaller in magnitude. These results suggest against an explanation that is driven by demand such as increased population growth, which we would expect to affect all types of physicians. While emergency physicians are also obviously tied to emergency departments and new hospitals we also do not find changes in concentration for other physician types like general surgeons or internal medicine physicians who would also likely concentrate in response to new hospitals.

Instead, our results suggest a supply-sided explanation that allows for concentration of physicians who are trained as family physicians. This may relate to the portability of the degree that CCFP(EM) and family physicians both have. Whereas the other physician types in our sample usually work in hospital settings, family physicians can work in outpatient settings^20^. This means they can more readily move to places that they prefer, such as urban settings. In the case of Canada where supply of healthcare is often highly constrained and demand for care is high^15^, this portability might mean these physicians can access improved urban amenities without sacrificing income.

Our findings have policy implications as they provide some insight into whether relying on less extensively trained emergency care providers may be effective at improving equity. Our results suggest that in the case of Ontario, relying on delivery of emergency care through this avenue of less extensively trained physicians has not improved equity over time. This has some implications for improving access to emergency care through physician extenders as these may not solve supply issues in rural areas. This suggests that strategy to deliver care into underserved areas requires more nuance than only relying on reduced training length.

### 4.1. Limitations and future research

Our approach does have several limitations. First, while Ontario provides several useful aspects, it could be limited in its external validity. Ontario has a single payer health insurance system and care is, to a large extent, centrally managed. The time period we examine is subject to major policy changes both in terms of health supply and health demand. For example, the COVID-19 pandemic occurred over our period of observation. There was also a significant increase in population due to immigration during this period, especially around southern Ontario^14^. We are limited in our ability to evaluate whether population in particular is driving our results. Our results might also be correlated with changes in locations of hospitals or EDs. However, we note that we would expect population changes or changes in hospital location to drive concentration of other physician types as well. We do not find similar large-scale changes in Moran’s I estimated for these alternate groups of physicians. However, policy may also be affecting our estimates in a way that only concentrates emergency medicine physicians. Most emergency care in Ontario is funded through volume-based hospital alternate payment plans and EM-specific jobs are one major driver of where these physicians practice relative to other specialties.

Finally, we note limitations related to estimating Moran’s I measures. These include issues related to the modifiable areal unit problem, sensitivity to the weighting matrix used, and stationarity. We have taken steps to avoid the modifiable areal unit problem through use of our tessellated map^17^ and our sensitivity checks in Table e2 also alter the spatial weighting matrix. However, one limitation that we do not stress test is stationarity across geographic units. It is likely that there are different spatial concentration relationships in northwest Ontario relative to southern Ontario which our models do not take into account. These would be a particularly interesting avenue for sensitivity analyses in future research.

## Supporting information

Supplementary Figure e1: Population Weighted Density of Physicians by Census FSA

Supplementary Figure e2: Local Morans I (raw) for CCFM(EM) and FRCPC specialists.

Supplementary Figure e3: Counts of Emergency Physicians within Ontario, tessellated square map.

Supplementary Figure e4: Counts of Emergency Physicians within Southern Ontario, tessellated square map.

## Data Availability

The authors can provide the aggregated data, as privacy allows, and the underlying code to replicate this project upon reasonable request to the authors.

## 5. Conclusion

We find increases in the geographic concentration of emergency physicians in the province of Ontario during the period of 2015 to 2024. Physicians have been concentrating to more urban areas in the province. This has occurred almost exclusively in less extensively trained CCFP(EM) providers.

## REFERENCES

1. Drummond AJ. No room at the inn: overcrowding in Ontario’s emergency departments. CJEM [Internet]. 2002 Mar [cited 2025Apr28];4(2):91–7. Available from: https://www.cambridge.org/core/product/identifier/S1481803500006187/type/journal_article

2. The Collaborative Working Group on the Future of Emergency Medicine in Canada (CWG-EM):, Sinclair D, Abu-Laban RB, Toth P, LeBlanc C, Eisener-Parsche P, et al. Emergency Medicine Training and Practice in Canada: Celebrating the Past & Evolving for the Future. CJEM [Internet]. 2017 Jul [cited 2025Apr28];19(S2):S1–8. Available from: https://www.cambridge.org/core/product/identifier/S1481803517003724/type/journal_article

3. Unprecedented and Worsening: Ontario’s Local Hospital Closures 2023 [Internet]. 2023 Dec. Available from: https://www.ontariohealthcoalition.ca/wp-content/uploads/final-report-hospital-closures-report.pdf

4. Bennett CL, Sullivan AF, Ginde AA, Rogers J, Espinola JA, Clay CE, et al. National Study of the Emergency Physician Workforce, 2020. Annals of Emergency Medicine [Internet]. 2020 Dec [cited 2025Apr28];76(6):695–708. Available from: https://linkinghub.elsevier.com/retrieve/pii/S0196064420305011

5. Gummerson S, Smith M, Warren O. Effect of an Emergency Department Closure on Homeless Patients and Adjacent Hospitals. Western Journal of Emergency Medicine [Internet]. 2022 Apr [cited 2025Apr28];23(3):368–74. Available from: https://escholarship.org/uc/item/4m76s4zh

6. Harrington DT, Connolly M, Biffl WL, Majercik SD, Cioffi WG. Transfer Times to Definitive Care Facilities Are Too Long: A Consequence of an Immature Trauma System. Annals of Surgery [Internet]. 2005 Jun [cited 2025Apr28];241(6):961–8. Available from: https://journals.lww.com/00000658-200506000-00013

7. Nolan B, Haas B, Tien H, Saskin R, Nathens A. Patient, Paramedic and Institutional Factors Associated with Delays in Interfacility Transport of Injured Patients by Air Ambulance. Prehospital Emergency Care [Internet]. 2020 Nov [cited 2025Apr28];24(6):793–9. Available from: https://www.tandfonline.com/doi/full/10.1080/10903127.2019.1701159

8. Randle T, Garg A, Mago V, Choudhury S, Ohle R, Strasser R, et al. Staffing rural emergency departments in Ontario: The who, what and where. Canadian Journal of Rural Medicine [Internet]. 2023 [cited 2025Apr28];28(2):73. Available from: https://journals.lww.com/10.4103/cjrm.cjrm_51_22

9. Patel SY, Auerbach D, Huskamp HA, Frakt A, Neprash H, Barnett ML, et al. Provision of evaluation and management visits by nurse practitioners and physician assistants in the USA from 2013 to 2019: cross-sectional time series study. BMJ [Internet]. 2023 Sep [cited 2025Apr28];:e73933. Available from: https://www.bmj.com/lookup/doi/10.1136/bmj-2022-073933

10. Steiner IP. Emergency medicine practice and training in Canada. CMAJ [Internet]. 2003 Jun [cited 2025Apr28];168(12):1549– 50. Available from: https://www.cmaj.ca/content/168/12/1549

11. Gregory Powell D. Canadian emergency medicine. The American Journal of Emergency Medicine [Internet]. 1984 Mar [cited 2025Apr28];2(2):171. Available from: https://linkinghub.elsevier.com/retrieve/pii/S0735675784800157

12. Elyas R. 31. The birth of a new specialty: The history of emergency medicine in Canada. Clinical & Investigative Medicine [Internet]. 2007 Aug [cited 2025Apr28];30(4):44. Available from: http://cimonline.ca/index.php/cim/article/view/2791

13. McEwen J, Borreman S, Caudle J, Chan T, Chochinov A, Christenson J, et al. Position Statement on Emergency Medicine Definitions from the Canadian Association of Emergency Physicians. CJEM [Internet]. 2018 Jul [cited 2025Apr28];20(4):501– 6. Available from: https://www.cambridge.org/core/product/identifier/S1481803518003767/type/journal_article

14. Shumanty R, Charbonneau P, Martel L. Canada’s Fastest Growing and Decreasing Municipalities from 2016 to 2021: Census of Population, 2021. 2022.

15. Gruber J. Financing Health Care Delivery. Annual Review of Financial Economics [Internet]. 2022 Nov [cited 2025Apr28];14(1):209–29. Available from: https://www.annualreviews.org/doi/10.1146/annurev-financial-111620-112740

16. Anselin L. Local Indicators of Spatial Association—LISA. Geographical Analysis [Internet]. 1995 Apr [cited 2025Apr28];27(2):93–115. Available from: https://onlinelibrary.wiley.com/doi/10.1111/j.1538-4632.1995.tb00338.x

17. Zhu H, Liu J, Zhou X, Chen X, Qiu X, Bello RL, et al. The Ontario Climate Data Portal, a user-friendly portal of Ontario-specific climate projections. Scientific Data [Internet]. 2020 May [cited 2025Apr28];7(1):147. Available from: https://www.nature.com/articles/s41597-020-0489-4

18. Atkinson P, McGeorge K, Innes G. Saving emergency medicine: is less more?. Canadian Journal of Emergency Medicine [Internet]. 2022 Jan [cited 2025Apr28];24(1):9–11. Available from: https://link.springer.com/10.1007/s43678-021-00237-1

19. Collier R. Canada’s emergency medicine shortfall. Canadian Medical Association Journal [Internet]. 2016 Aug [cited 2025Apr28];188(11):E246–6. Available from: http://www.cmaj.ca/lookup/doi/10.1503/cmaj.109-5292

20. Rabinowitz HK, Paynter NP. The Rural vs Urban Practice Decision. JAMA [Internet]. 2002 Jan [cited 2025Apr28];287(1):113. Available from: http://jama.jamanetwork.com/article.aspx?doi=10.1001/jama.287.1.113-JMS0102-7-1

