## Supplementary figures and images for "Do Faster-Trained Physicians Fill the Gaps? Geographic Concentration of Emergency Medicine physicians with different postgraduate training in Ontario Canada"

### Supplementary Figure e1: Population Weighted Density of Physicians by Census FSA

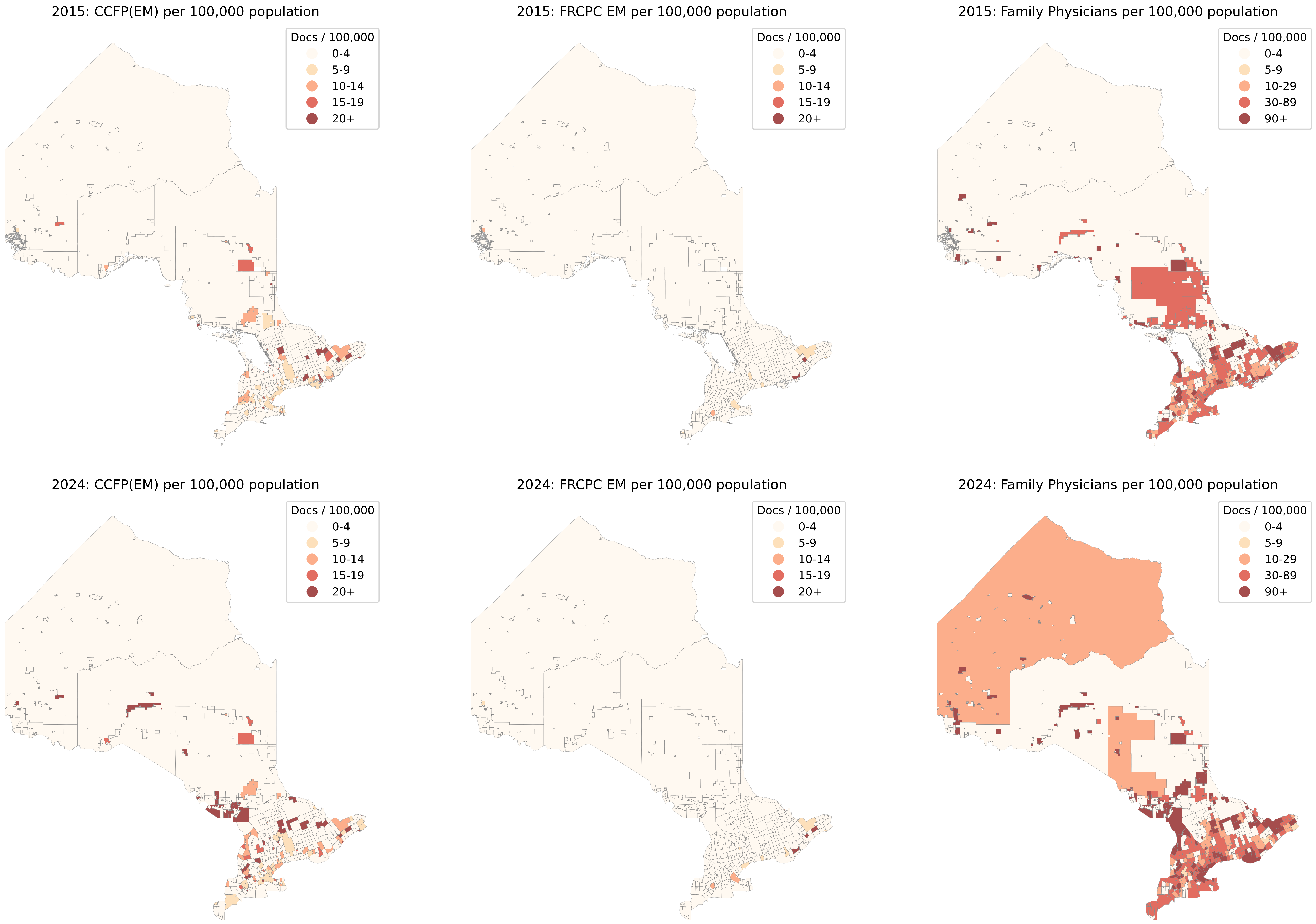

### Supplementary Figure e2: Local Morans I (raw) for CCFM(EM) and FRCPC specialists.

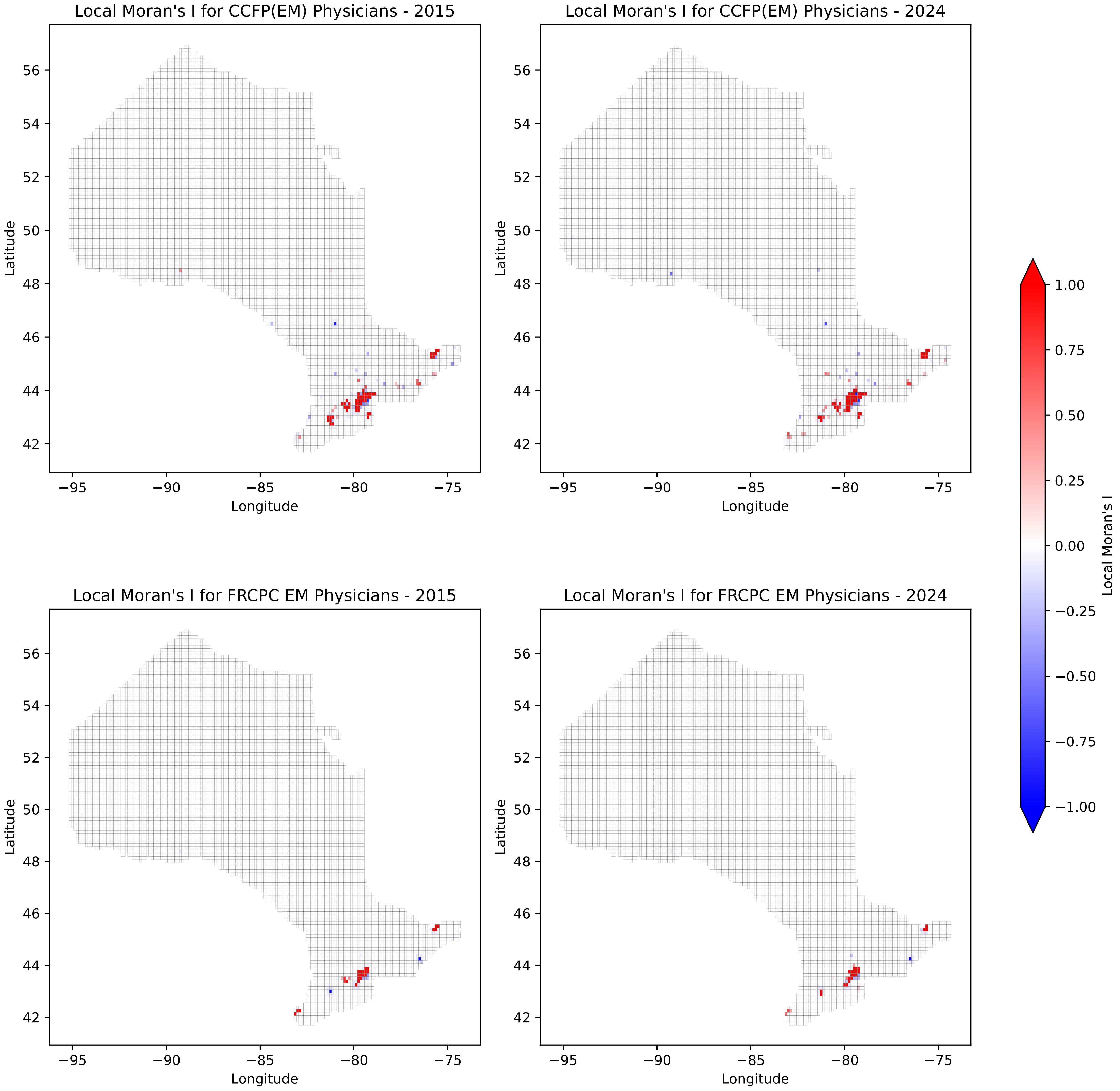

### Supplementary Figure e3: Counts of Emergency Physicians within Ontario, tessellated square map.

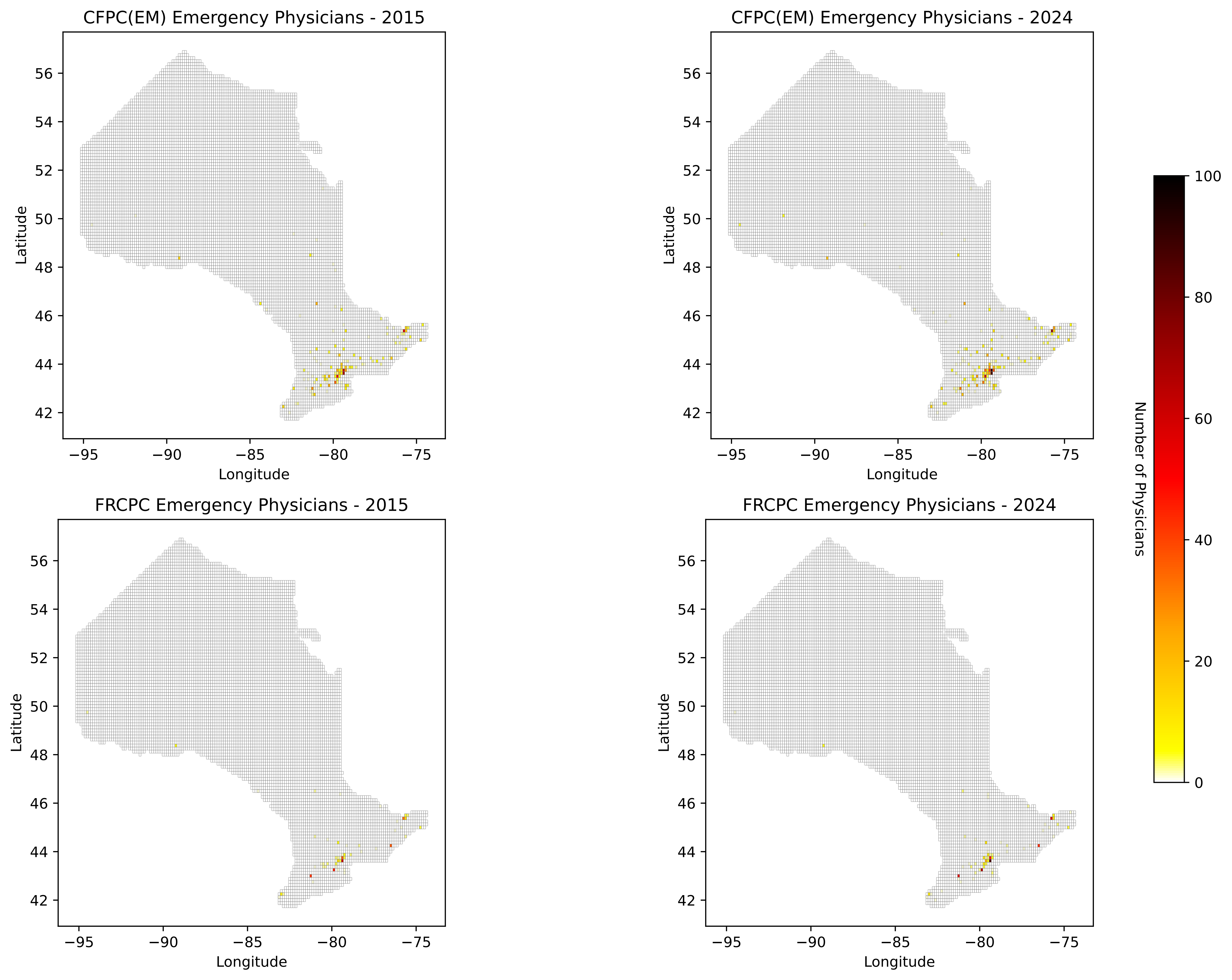

### Supplementary Figure e4: Counts of Emergency Physicians within Southern Ontario, tessellated square map.

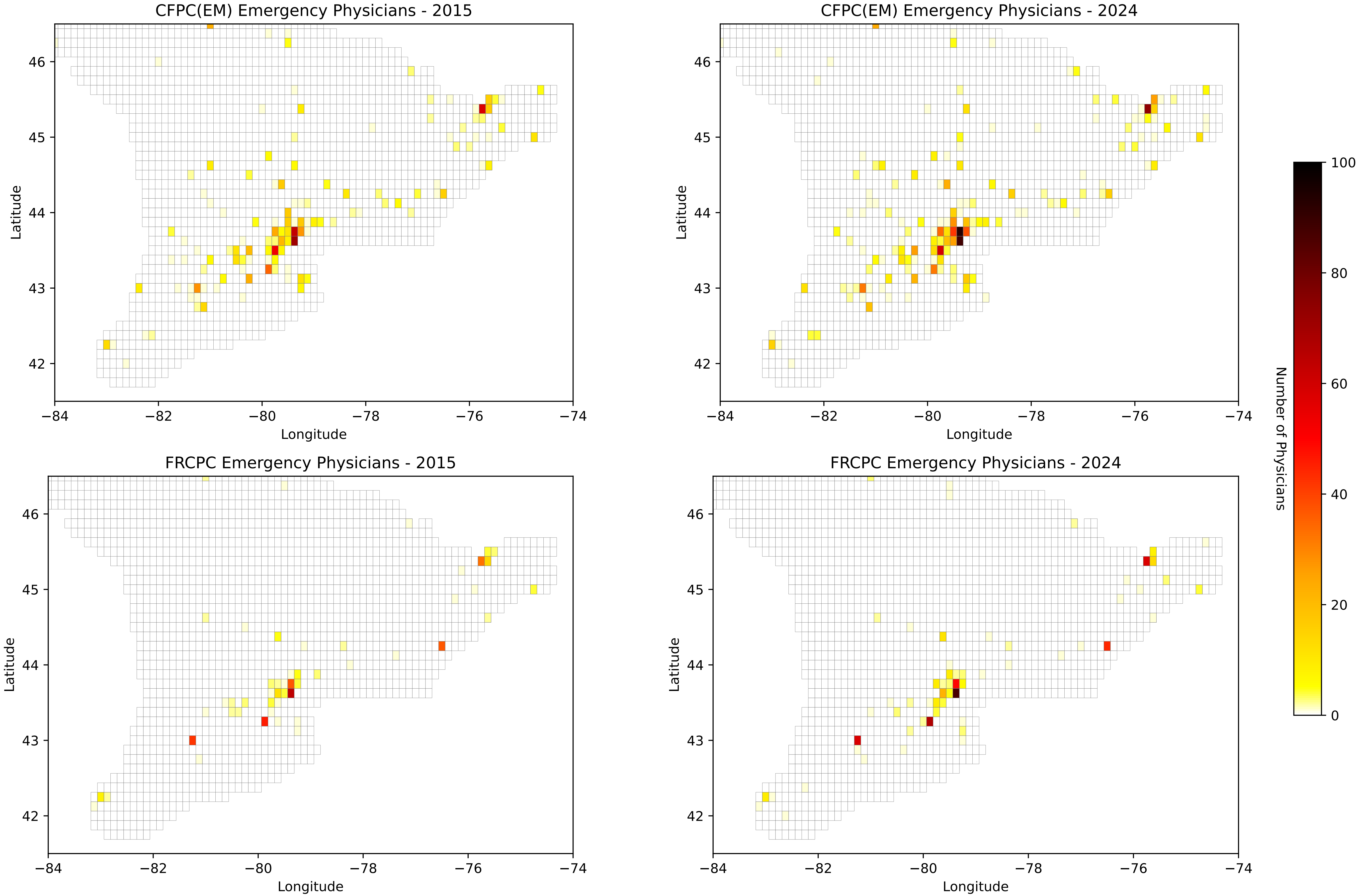
