## Supplementary Table e1 for "Do Faster-Trained Physicians Fill the Gaps? Geographic Concentration of Emergency Medicine physicians with different postgraduate training in Ontario Canada"

|  | **2015 - CCFP(EM)** | | | **2024 - CCFP(EM)** | | |
| --- | --- | --- | --- | --- | --- | --- |
|  | No specialist in Community | Specialist in Community | Total | No specialist in Community | Specialist in Community | Total |
| N | 286 (54.5%) | 239 (45.5%) | 525 (100.0%) | 291 (54.7%) | 241 (45.3%) | 532 (100.0%) |
| Agricultural production | 169.24 (395.15) | 234.39 (658.09) | 199.39 (533.66) | 151.98 (346.38) | 258.90 (657.32) | 201.23 (515.79) |
| Average age | 41.23 (3.55) | 41.80 (3.14) | 41.49 (3.38) | 41.81 (5.95) | 42.28 (4.37) | 42.02 (5.28) |
| Employed | 10428.80 (8201.57) | 15797.17 (10042.33) | 12913.77 (9477.50) | 9876.21 (7467.39) | 15490.38 (10273.02) | 12462.39 (9293.93) |
| Median income | 37054.25 (8370.21) | 34374.59 (7507.07) | 35813.86 (8085.97) | 43765.48 (9443.23) | 41712.50 (7220.58) | 42819.77 (8545.71) |
| Population | 20452.74 (16239.28) | 32097.44 (19660.78) | 25762.01 (18782.57) | 21552.54 (15727.44) | 34031.99 (21375.94) | 27301.23 (19544.04) |
|  | **2015 - FRCPC EM** | | | **2024 - FRCPC EM** | | |
|  | No specialist in Community | Specialist in Community | Total | No specialist in Community | Specialist in Community | Total |
| N | 440 (83.8%) | 85 (16.2%) | 525 (100.0%) | 436 (82.0%) | 96 (18.0%) | 532 (100.0%) |
| Agricultural production | 213.64 (574.32) | 127.82 (231.28) | 199.39 (533.66) | 207.94 (545.23) | 171.51 (358.19) | 201.23 (515.79) |
| Average age | 41.47 (3.41) | 41.63 (3.24) | 41.49 (3.38) | 41.91 (5.60) | 42.53 (3.50) | 42.02 (5.28) |
| Employed | 12232.93 (9103.63) | 16334.00 (10579.69) | 12913.77 (9477.50) | 11868.01 (9053.59) | 15093.75 (9919.16) | 12462.39 (9293.93) |
| Median income | 35998.06 (8079.80) | 34888.53 (8101.14) | 35813.86 (8085.97) | 42890.12 (8835.16) | 42508.33 (7157.06) | 42819.77 (8545.71) |
| Population | 24332.13 (18150.77) | 33113.28 (20316.97) | 25762.01 (18782.57) | 25981.70 (19148.47) | 33142.90 (20298.47) | 27301.23 (19544.04) |

Table e1: Summary statistics by FSAs that have MDs by year and physician type. Standard deviations are in parentheses, except for N where the parenthetical is a percentage of the total.
