## Supplementary Table e2 for "Do Faster-Trained Physicians Fill the Gaps? Geographic Concentration of Emergency Medicine physicians with different postgraduate training in Ontario Canada"

| Weight | 19km Distance Band | | | | 38km Distance Band | | | | 96km Distance Band | | | |
| --- | --- | --- | --- | --- | --- | --- | --- | --- | --- | --- | --- | --- |
| Year | 2015 | | 2024 | | 2015 | | 2024 | | 2015 | | 2024 | |
| Specialty | Moran's I | p | Moran's I | p | Moran's I | p | Moran's I | p | Moran's I | p | Moran's I | p |
| CCFP Emergency Medicine | 0.2343 | 0.001 | 0.3083 | 0.001 | 0.1482 | 0.001 | 0.1756 | 0.001 | 0.0758 | 0.001 | 0.0834 | 0.001 |
| FRCPC Emergency Medicine | 0.0955 | 0.001 | 0.1028 | 0.001 | 0.0425 | 0.001 | 0.0523 | 0.001 | 0.0216 | 0.001 | 0.0249 | 0.001 |
| Emergency Medicine - All Types | 0.1894 | 0.001 | 0.2396 | 0.001 | 0.1124 | 0.001 | 0.1307 | 0.001 | 0.0557 | 0.001 | 0.0613 | 0.001 |
| Any Non-EM Physician | 0.2614 | 0.001 | 0.3063 | 0.001 | 0.1342 | 0.001 | 0.1654 | 0.001 | 0.0613 | 0.001 | 0.0758 | 0.001 |
| Family Medicine | 0.3784 | 0.001 | 0.404 | 0.001 | 0.22 | 0.001 | 0.2376 | 0.001 | 0.1082 | 0.001 | 0.115 | 0.001 |
| General Surgery | 0.2203 | 0.001 | 0.2305 | 0.001 | 0.1196 | 0.001 | 0.1282 | 0.001 | 0.0617 | 0.001 | 0.0667 | 0.001 |
| Internal Medicine | 0.2078 | 0.001 | 0.2331 | 0.001 | 0.1082 | 0.001 | 0.1279 | 0.001 | 0.051 | 0.001 | 0.0589 | 0.001 |
| Pediatrics | 0.1566 | 0.001 | 0.1613 | 0.001 | 0.076 | 0.001 | 0.079 | 0.001 | 0.0324 | 0.001 | 0.0353 | 0.001 |

Table e2: Estimates of Moran’s I by specialty and year. Our main estimates use a 19km distance weighting matrix. We also demonstrate sensitivity to this choice by altering the distance band. Higher estimates of Moran’s I means higher concentration of physicians ie. tessellated squares with high numbers of physicians are more likely to be close to other tessellated squares with high numbers of physicians. p values simulated using a permutations approach.
